# ASSAY OF THE DREEM DEVICE ON SLEEP METRICS AND AN EXPLORATION OF SLEEP STAGING IN CHRONIC SHORT SLEEPERS DURING TIME IN BED EXTENSION

**DOI:** 10.1101/2023.06.27.23291956

**Authors:** Zachary Mallender, Christopher M. Depner

**Affiliations:** Department of Health and Kinesiology, University of Utah, Salt Lake City, Utah, USA

## Abstract

Despite clear research findings showing that sleeping less than seven hours per night has an array of health consequences, over 1 in 3 American adults report sleeping less than seven hours per night. Many studies exploring the consequences of insufficient sleep are restricted to small sample sizes and are of relatively short duration due to a significant cost of gold-standard polysomnography in terms of participant burden, expense, time, and reliance on trained sleep technicians. Additionally, many studies of short sleep duration use a paradigm of experimental sleep restriction on otherwise healthy sleepers, which excludes people who chronically obtain short sleep duration over months to years. Here, we explore possible solutions to these issues by implementing a sleep extension protocol in 14 adults (average age 20.6±2.5y; +/- SD) with self-reported habitual sleep duration less than 6.5h/night. Participants completed 2 weeks of baseline monitoring (habitual short sleep duration) and then were instructed to increase time in bed to ≥8h/night for four weeks. Sleep was monitored using wrist-actigraphy and the Dreem 2 headband, a wireless dry electrode consumer electroencephalography (EEG) device. Compared to wrist-actigraphy, the Dreem 2 shows minimal systemic skew for nights with data quality over 75% (as assigned by the Dreem algorithm). However, Bland Altman analysis shows significant random error with limits of agreement approximately +/- 70 minutes between actigraphy and the Dreem. Exploration of sleep metrics from the Dreem 2 during baseline short sleep versus sleep extension revealed an increase in total sleep time; increase in all recorded sleep stages; and no significant changes in sleep onset latency, wakefulness after sleep onset, or sleep efficiency. Although several limitations of producing high quality data were identified, the Dreem 2 headband shows promise as a home environment sleep research device. With an improvement in data quality, the Dreem headband, or another wireless consumer sleep device, has the potential to help advance the sleep field in ways that were previously inaccessible with clinical PSG.

## INTRODUCTION

Approximately 35% of American adults report sleeping less than 7 hours per night, the minimum recommended by the Sleep Research Society and American Academy of Sleep Medicine [1, 2]. Insufficient sleep is a risk factor for many health issues including diabetes, obesity, poor mental health, and impaired cognitive performance [1-5]. Typically, sleep research in the laboratory is based on the paradigm of experimentally restricting sleep in healthy young adults to 4-5 hours of sleep per night for 1-14 nights [6,7], followed by ∼2-7 days of recovery sleep to relieve the sleep restriction [8]. This paradigm allows precise analyses of the physiological health consequences of experimental sleep restriction in otherwise healthy adults. However, this type of sleep research is restricted in scope, both in study duration and in participant number, due to the cost and burden of labor-intensive sleep restriction where participants live in the laboratory 24 hours per day. It also unclear if research findings from experimental sleep restriction directly translate to people with habitual short sleep duration in the free-living environment. Because it is not feasible or ethical to conduct months-long studies of experimental sleep restriction outside the laboratory, there is a need for noninvasive large-scale at home sleep research. Such a research approach has large potential to advance the entire field of sleep research. Importantly, studying sleep in the home-environment is likely more ecologically valid compared to experimentally restricting sleep in the laboratory setting.

Polysomnography (PSG) is the currently accepted gold standard of in-laboratory and clinical sleep monitoring. PSG requires the application of electroencephalography (EEG) electrodes to the scalp, electromyography (EMG) electrodes to the facial muscles and limbs, electrooculography (EOG), electrocardiography (EKG), pulse oximetry, thoracic and abdominal belts, and nasal (or oro-nasal) flow sensors to measure breathing patterns [9]. EEG data of brain activity is used to determine the stage of sleep that the participant is in. Stages are broadly represented as N1, N2, N3 (deep sleep), and REM (rapid eye movement) sleep. EEG can also be used as a measure of homeostatic sleep pressure using delta power (0.5-4.5 hertz), which is a measure of slow wave activity (SWA). SWA is thought to represent homeostatic sleep pressure due to observations of increased SWA during restricted sleep and decreased SWA during recovery sleep in laboratory settings [10].

Despite its strengths, PSG is expensive, requires technical expertise, and has a high participant burden as it involves applying 20-50 electrodes in precise locations on the head. All of these factors make long-term sleep monitoring in the home environment by PSG unfeasible, especially over time frames longer than a week. Ambulatory dry EEG devices offer a possible solution to this issue, combining some aspects of PSG with a more convenient and user-friendly design. This combination has the potential to allow for high quality EEG data to be gathered without the financial and technical limitations of PSG, and has the potential to significantly advance sleep research.

The Dreem 2 headband is one such dry-electrode consumer EEG device [11]. Preliminary data show the Dreem 2 headband provides sufficient quality sleep recordings and that the algorithm provided by Dreem is as accurate as certified Sleep Technicians scoring in-laboratory PSG data [13-15]. It should be noted however that most of this data is derived from studies funded by the Dreem company and these data are derived from overnight studies in the laboratory as opposed to the free-living home environment where the participant has full control responsibility over setting up the Dreem headband.

As such, we conducted a pilot study with the goal of exploring the quality of data recorded by the Dreem 2 headband device in a hands-off, at home environment. As we are broadly interested in the health consequences of habitual short sleep duration, we focus our analyses on participants who report habitually sleeping less than 6.5 hours a night. Analyses of this pilot study are part of a larger ongoing study that involves two weeks of baseline assessments followed by a 4-week sleep extension intervention with sleep monitored by wrist-actigraphy and the Dreem 2 headband in the home environment.

## METHODS

### Participants

The parent study is an ongoing, longitudinal, single-arm experimental protocol designed to examine metabolic changes in response to sleep extension in adults aged 18- 35 years who report sleeping less than 6.5 hours per night. All study procedures were approved by the University of Utah Institutional Review Board and all participants signed a written informed consent document prior to completing any study procedures. Inclusion criteria were: (1) men and women aged 18-35 years; (2) self-reported habitual sleep duration less than 6.5h/night; (3) normal body mass index of 18.5-24.9 kg/m^2^; (4) must have lived at Salt Lake City, UT, altitude or higher for at least three months. Exclusion criteria were: (1) Any clinically significant unstable medical or surgical condition within the last year (treated or untreated). (2) Any clinically significant psychiatric condition, as defined by DSM-IV-TR. I (3) Any clinically significant sleep disorder. (4) Use of prescription medications/supplements within one month or need of these medications at any time during the study. (5) Symptoms of active illness (e.g., fever). (6) Uncorrected visual impairment (7) History of shift work in prior year or travel more than one time zone in three weeks prior to study. (8) Participants must be entirely drug-free of illicit drugs, medications, nicotine and herbal products for one month prior to study. (9) Blood donation in the 30 days prior to inpatient study. (10) Ovulating women were selected on the basis of a history of regular menstrual cycle ranging in length from 25-32 days with a maximum of three days variation month-to-month. They had no history of prior gynecological pathology, were at least 1 year post-partum, not breast-feeding and not pregnant (HCG pregnancy test at screening and upon admission to the inpatient protocol).

Clinical overnight sleep disorders screening ensured participants did not have sleep disorders. Participants were screened for self-report medical disorders and medical screening with urine toxicology, complete blood count, and comprehensive metabolic panel helped ensure participants did not meet diagnostic criteria for medical disorders. Fifteen participants have completed the Dreem 2 portion of the study. One participant was removed from our current analyses due to an inability to wear the Dreem headband.

### Protocol

The complete protocol consisted of a two-week baseline monitoring segment followed by a four-week sleep extension segment. Each study segment was completed in the participant’s home environment. At baseline, participants were instructed to maintain their habitual sleep schedules. For sleep extension, participants were instructed to extend their time in bed to ≥8h/night. This target of 8h/night of time in bed was selected with the goal of having participants achieve the recommended 7h of nightly total sleep time [1,2]. Sleep was monitored throughout the study by wrist-actigraphy (Actiwatch Spectrum Plus; Philips Inc.) worn on their non-dominant hand and daily written sleep diaries [12]. During the final week of baseline and sleep extension segments sleep was also monitored using the Dreem 2 headband (study weeks 2 and 6) in the participants home environment.

### Sleep Extension Intervention

The study team worked with each participant to set target bed and waketimes that fit individual participant schedules with the goal of achieving 8 hours of time-in-bed per night. General sleep tips were also provided to participants including keeping a regular sleep and wake schedule on work-days and free days, avoiding caffeine in the afternoon, keep the bedroom environment cool and dark, and to minimize light exposure from electronic devices in the hour prior to bedtime. Participants sleep was also tracked daily in real-time by electronic sleep log. If a participant failed to completed the electronic sleep log for two consecutive days or was failing to extend their time in bed a member of the research team reached out to trouble shoot barriers. At the two-week point of the four-week sleep extension participants had an in-person study visit to review their actigraphy data, adjust target bed and waketimes if needed, and troubleshoot any barriers to sleep extension. One week of EEG data was collected similarly to the baseline segment with the Dreem 2 headband recording days 35-42 in the participants home environment.

### Dreem 2 Headband

The Dreem 2 headband is composed of a mix of flexible plastic panels and soft fabric headband, with an elastic strap mounted in the back [13]. Battery life is roughly 12 hours. It records data through 6 EEG electrodes, with 4 dry silicone rubber electrodes along the forehead and 2 proprietary electrodes positioned at the base of the back of the skull. This data, combined with an accelerometer and a pulse sensor, are initially assigned a quality from 0-100 by the onboard Dreem computer. The computer then takes the two highest quality EEG channels and uses a proprietary Dreem developed deep learning algorithm to automatically assign a sleep stage to each 30s epoch. Each night and each EEG channel within each night are assigned a quality score at the end of the recording based on the estimated percent of the recording that is scorable. Quality over 85% is listed as green, quality between 85%-70% is listed as yellow, and quality below 70% is listed as red. Data is uploaded to Amazon A3 servers owned by Dreem, where it is stored and can be accessed.

Participants were given initial in-person instructions, verbal and written, on how to wear the Dreem headband most effectively. Dreem data quality were monitored daily and participants were sent a follow-up text message if initial data had low quality as calculated automatically by the Dreem algorithm. Participants were instructed to wear the Dreem headband nightly for 7 days unless the headband was noticeably disruptive to their sleep.

### Bland-Altman Plots

To compare nightly total sleep time measured by the Dreem 2 versus wrist-actigraphy we used Bland-Altman plots. Dreem and actigraphy data were processed through R using a data package (BAplot.R, R version 4.2.1) for the production of Bland-Altman plots [14]. We assigned the Actiwatch data as the reference data and the Dreem 2 as experimental data. Data was processed using bootstrap to account for the low n value, and data was log transformed to minimize possible impacts of proportional bias of sleep duration, consistent with current recommendations [14].

### Exploratory Power Spectral Analysis

Following our initial determination that Dreem quality over 70% was sufficient for further data analysis, we attempted to compare nightly SWA in the baseline versus sleep extension study segments. Given the exploratory nature of this analysis we determined that a minimum of 4 nights of 70% or higher quality data within a single participant for both study segments was necessary. Because only three participants satisfied this metric, we determined that a case study of those three participants would be most useful in determining our ability to effectively perform a SWA analysis. To ensure spatial continuity of SWA, we restricted these case study analyses to one channel per participant, either F7-O1 or F8-O2. We used BrainVision Analyzer 2 software to process the Dreem EEG data (.EDF format), with a raw data analysis using filters for gradient (50μV/ms max), amplitude (minimum of −200 μV and a maximum of 200 μV), and low activity (minimum activity max-min of 0.5 μV). Only the first three NREM episodes of each night were analyzed to ensure the same number of sleep cycles were included in each night of data, consistent with other studies in the field [15]. The beginning of a NREM episode was defined as 2 consecutive 30 second epochs of N1 sleep, or one epoch of N2 sleep. A NREM episode ended upon one epoch of REM sleep, and a second NREM episode was not considered until at least 15 minutes had elapsed between REM episodes. The raw data inspection tool in Analyzer 2 was used to remove artifacts from the data, removing an average of 25.4% of the total data. We then performed a Fast Fourier Transformation of average delta power (0.5-4.5 hertz) to quantify SWA.

### Statistical analysis

All statistical processing was performed in R (v4.2.1) using Lme4 and Multcomp packages. Models were made using linear mixed effects models (LMER) to compare total sleep time for baseline and sleep extension conditions, using subject ID as a random variable to account for individual subject effects.

## RESULTS

Fourteen participants, 8 male and 6 female, completed the Dreem 2 headband portion of our study. Their age was 20.6±2.5y (mean±SD), with an average BMI of 22.6±2.28 (at study enrollment). Data was collected between 8/5/2021 and 11/12/2022.

**Figure 1:**
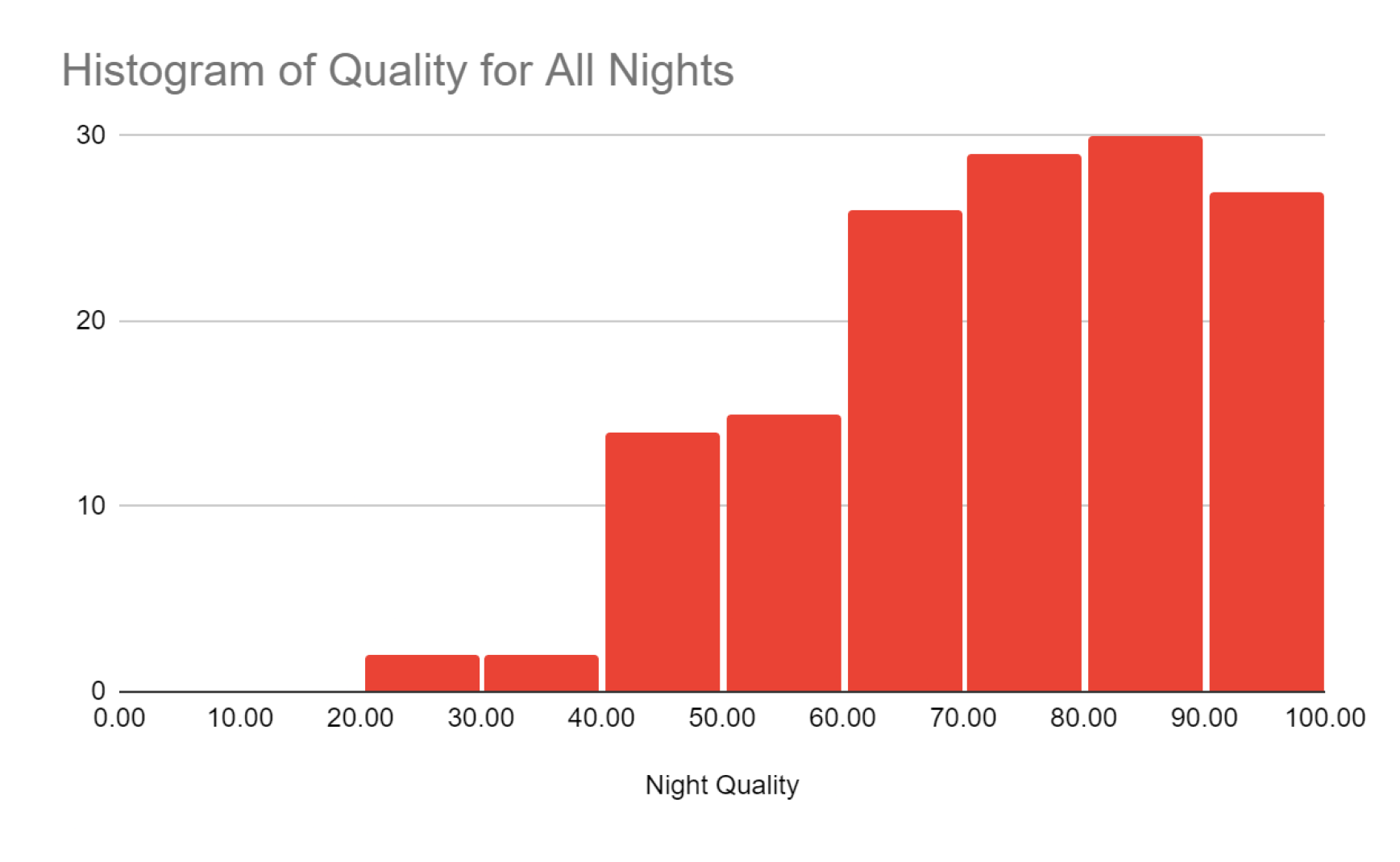
Histogram of Dreem assigned recording quality for all recorded nights. Each bin represents a 10% quality bracket from 0% to 100% quality. Dreem defines the quality metric as the percentage of the recording that could be effectively scored by a sleep technician.

### Comparing Total Sleep Time (TST) by Actiwatch and Dreem 2

Figure 2 represents all nights of Dreem 2 data tabulated by recording quality in 10% bins. When all Dreem 2 data, regardless of quality score, were included in the analyses (Figure 2A) there was significant heteroscedastic bias with a negative skew such that TST tended to be lower for the Dreem 2 versus Actiwatch when the Actiwatch TST was longer than ∼450 minutes per night, but TST tended to be higher for the Dreem 2 versus Actiwatch when the Actiwatch TST was shorter than ∼250 minutes. Interestingly there is little bias between the mean TST of the devices, possibly implying that a portion of the negative skew is introduced by outliers above 450 minutes of sleep. When only nights with Dreem 2 quality scores >85% were included in the analyses (Figure 2B) there was no significant bias or difference in mean TST between Dreem 2 and Actiwatch. Similarly, when only nights with Dreem 2quality scores >75% were included in the analyses (Figure 2C) there was no significant bias or difference in mean TST between Dreem 2 and Actiwatch. When only nights with Dreem 2quality scores <70% were included in the analyses (Figure 2D) there was significant heteroscedastic bias with a negative skew such that TST tended to be lower for the Dreem 2 versus Actiwatch when the Actiwatch TST was longer than ∼450 minutes per night, but TST tended to be higher for the Dreem 2 versus Actiwatch when the Actiwatch TST was shorter than ∼250 minutes. This is reflected by the significant negative bias of the mean TST. Additionally, regardless of the nightly quality score stratification, all comparisons showed the LOA were wider as TST measured by the Actiwatch was longer.

**Figure 2:**
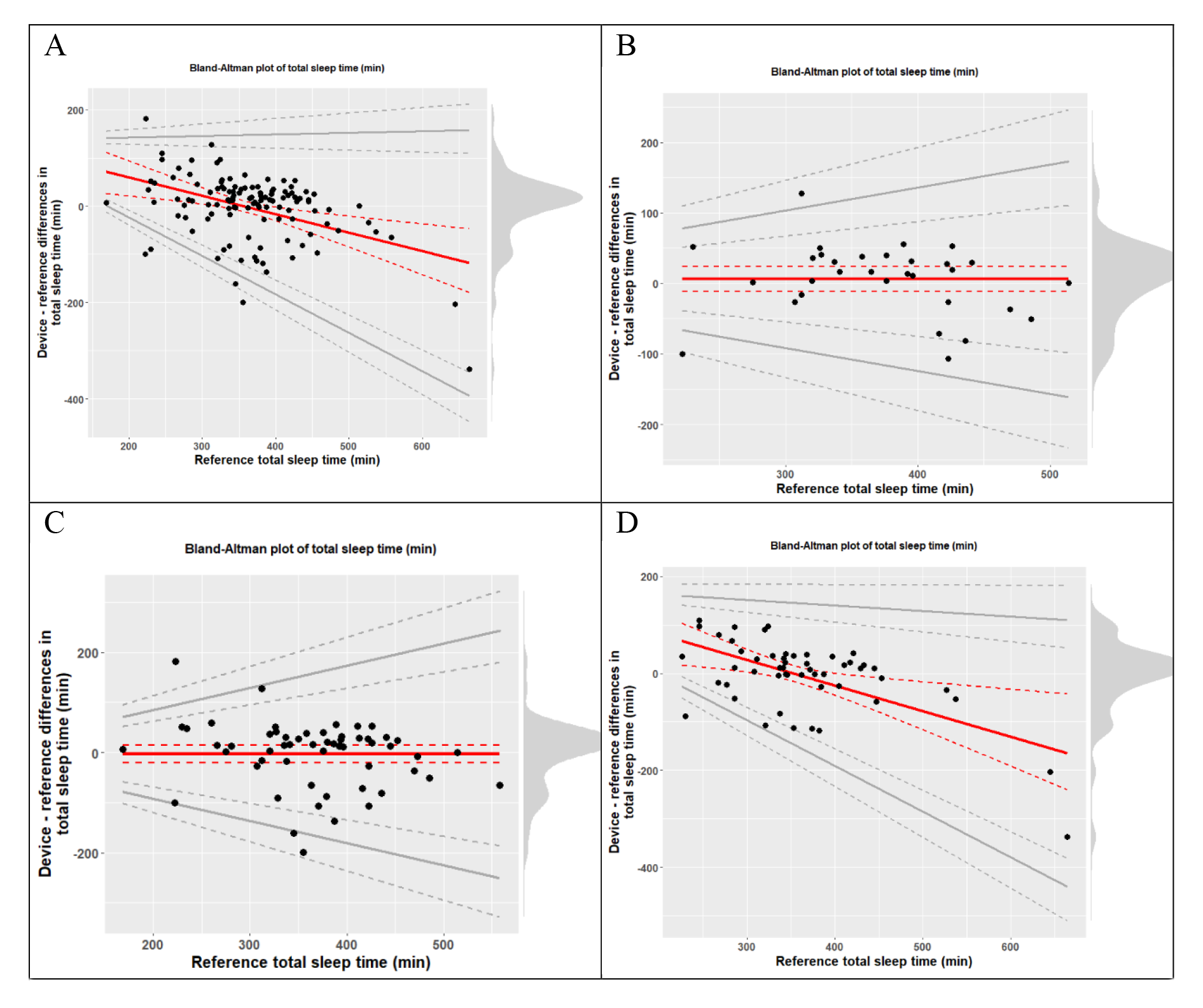
Bland-Altman plots showing comparative measure of total sleep time in minutes. Actiwatch data served as a reference device, which Dreem data was compared against. The red line represents calculated overall skew, and the solid grey lines represent the limits of agreement (LOA) for the two data sets. Dotted lines represent respective 95% CI. Panel A: all nights of data collected. Panel B: nights of greater than 85% quality. Panel C: nights of greater than 75% quality. Panel D: nights of less than 70% quality. Systemic skew underreporting sleep time for longer sleep episodes can be clearly seen in lower quality data, but is not present in green quality data. All data has roughly similar LOA, representing overall agreement of the Actiwatch and the Dreem headband, with a small increase in LOA for lower quality data. Data of 75% or higher virtually eliminates systemic skew.

When analyses were limited to only high-quality data from Dreem 2 (more than 85% scorable), there was minimal systemic error between the Dreem 2 data and wrist-actigraphy data. However, the limits of agreement for all plots were broad, with good quality LOA falling ∼65 minutes above or below zero, meaning that 95% of additional trials would fall within an approximately two-hour window. This indicates that while the Dreem 2 headband seems accurate for high quality data, it still lacks precision.

**Table 1:**
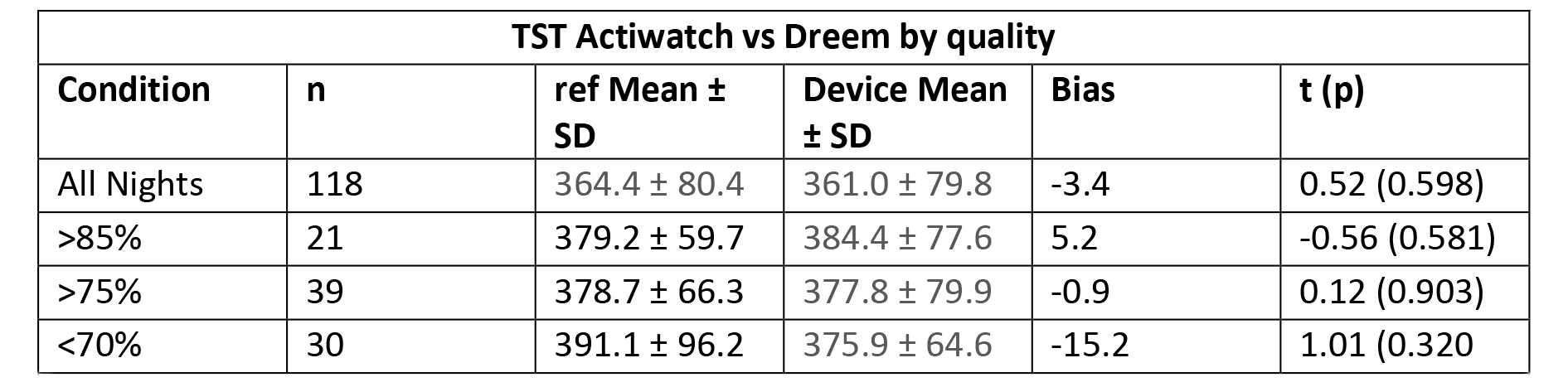
Reference (Actiwatch) and device (Dreem 2 headband) mean and standard deviation for TST. Bias is calculated by the difference between means, with negative bias suggesting device underreporting and positive bias suggesting device over-reporting. Columns are organized by quality, determined by the onboard Dreem algorithm. T values were calculated using a paired T-test. Ref, reference.

### Dreem 2 Sleep Stage Data during Baseline and Sleep Extension

Next, we analyzed TST and sleep staging measures for the baseline and sleep extension segments. Based on the generated Bland-Altman plots we determined that using data of 75% quality or higher allowed for a balance of including enough data while still eliminating systemic skew due to low quality data (Table 2). Using all nights of Dreem 2 data our results show participants achieved sleep extension with a mean of 5.5±0.2 hours of TST at baseline and significantly increased (p < 0.001) TST of 6.8±0.2 hours during sleep extension. An analysis of data of all qualities is included for reference (Table 3). The results of our linear mixed-model analysis for TST showed an intercept of 308±14.7 minutes (Estimate ± Standard error), with an increased TST by ∼101 minutes (standard error 14.5 minutes, p value of <0.001) during sleep extension.

**Table 2:**
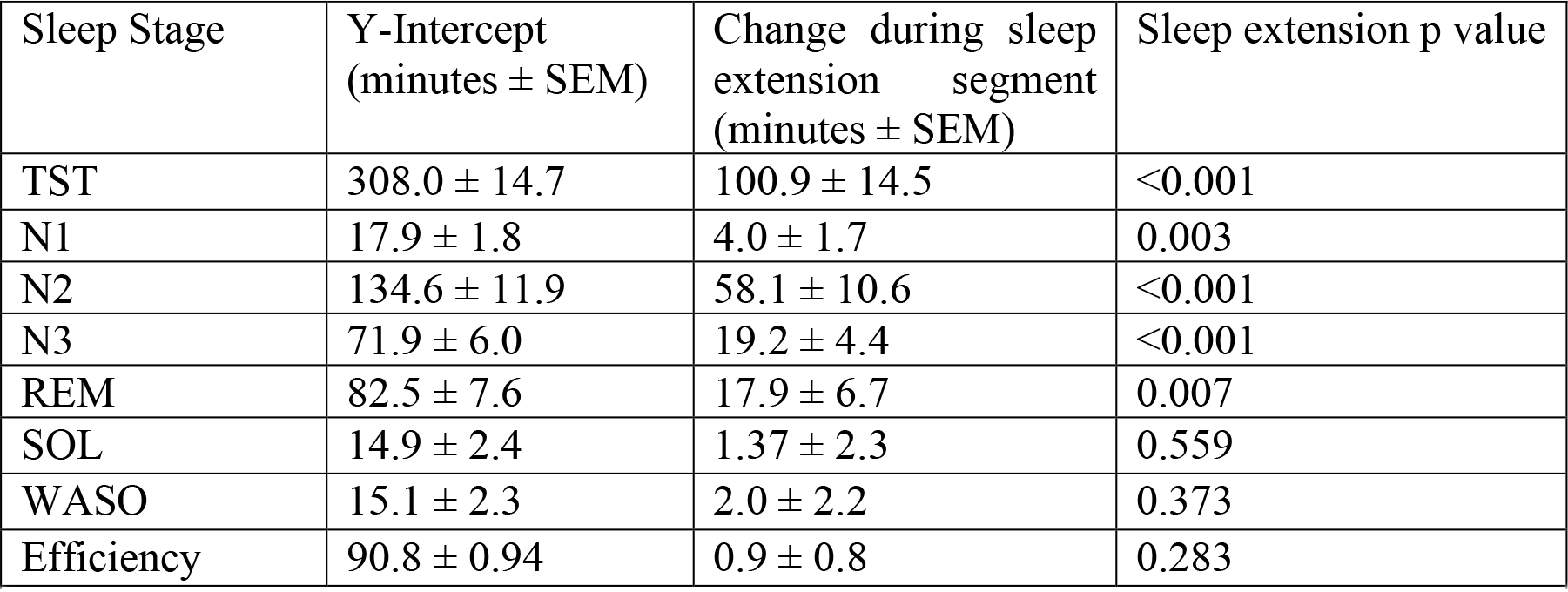
LMER of individual sleep stages as reported by the Dreem scoring algorithm for nights of >75% quality.

**Table 3:** LMER of individual sleep stages for all nights of all quality

| Sleep Stage | Y-Intercept<br>(minutes ± SEM) | Change during sleep<br>extension segment<br>(minutes ± SEM) | Sleep extension p<br>value |
| --- | --- | --- | --- |
| TST | $324.6 \pm 11.1$ | $83.3 \pm 11.6$ | $<0.001$ |
| N1 | $17.4 \pm 1.5$ | $5.8 \pm 1.2$ | $<0.001$ |
| N2 | $142.2 \pm 10.6$ | $46.7 \pm 7.5$ | $<0.001$ |
| N3 | $76.6 \pm 5.2$ | $7.2 \pm 4.2$ | 0.085 |
| REM | $87.5 \pm 6.4$ | $23.9 \pm 5.4$ | $<0.001$ |
| SOL | $21.0 \pm 4.7$ | $-2.4 \pm 6.1$ | 0.699 |
| WASO | $17.5 \pm 2.8$ | $8.2 \pm 2.7$ | 0.002 |
| Efficiency | $90.7 \pm 0.9$ | $-1.3 \pm 0.8$ | 0.091 |
| Table 3: LMER of individual sleep stages for all nights of all quality |  |  |  |

Based on the Dreem 2 automatic sleep staging algorithm, the duration of all sleep stages (N1, N2, N3, REM) increased during sleep extension versus baseline. Based on average change in minutes, stage N1 sleep increased the least and stage N2 increased the most. We did not detect significant changes in sleep onset latency (SOL) (p=0.559), sleep efficiency (SE) (p=0.283), or wakefulness after sleep onset (WASO) (p=0.373).

### Exploratory Delta Power Analysis

Finally, we performed a proof-of-concept case study to help inform the feasibility of using the Dreem 2 data for power spectral analyses when data collection is conducted in the home environment without supervision from the research team (Figure 3). We selected the three participants with the highest overall data quality for baseline and sleep extension, and used only nights with >70% total quality based on the Dreem algorithm. As previously noted, we limited these analyses to one channel per participant, either F7- O1 or F8-O2. Interestingly, we found that overall nightly data quality did not consistently reflect individual channel quality. Specifically, only 23% of the nights with an overall quality >70% also had a nightly channel quality with >70% for either the F7-O1 or F8- O2 channels. Furthermore, our data processing workflow (described in Methods) for this step flagged over ∼25% of the data as artifact on average, with a maximum removal of 56.5% and a minimum removal of 5.5% per night. With our small sample size and relatively low level of overall data quality, compared to overnight clinical PSG in a sleep lab, we cannot draw strong conclusions from calculated SWA. Nonetheless, two participants showed a SWA increase of approximately one μV^2 during extension versus baseline, and the third showed a decrease by one μV^2. During extension versus baseline.

**Figure 3:**
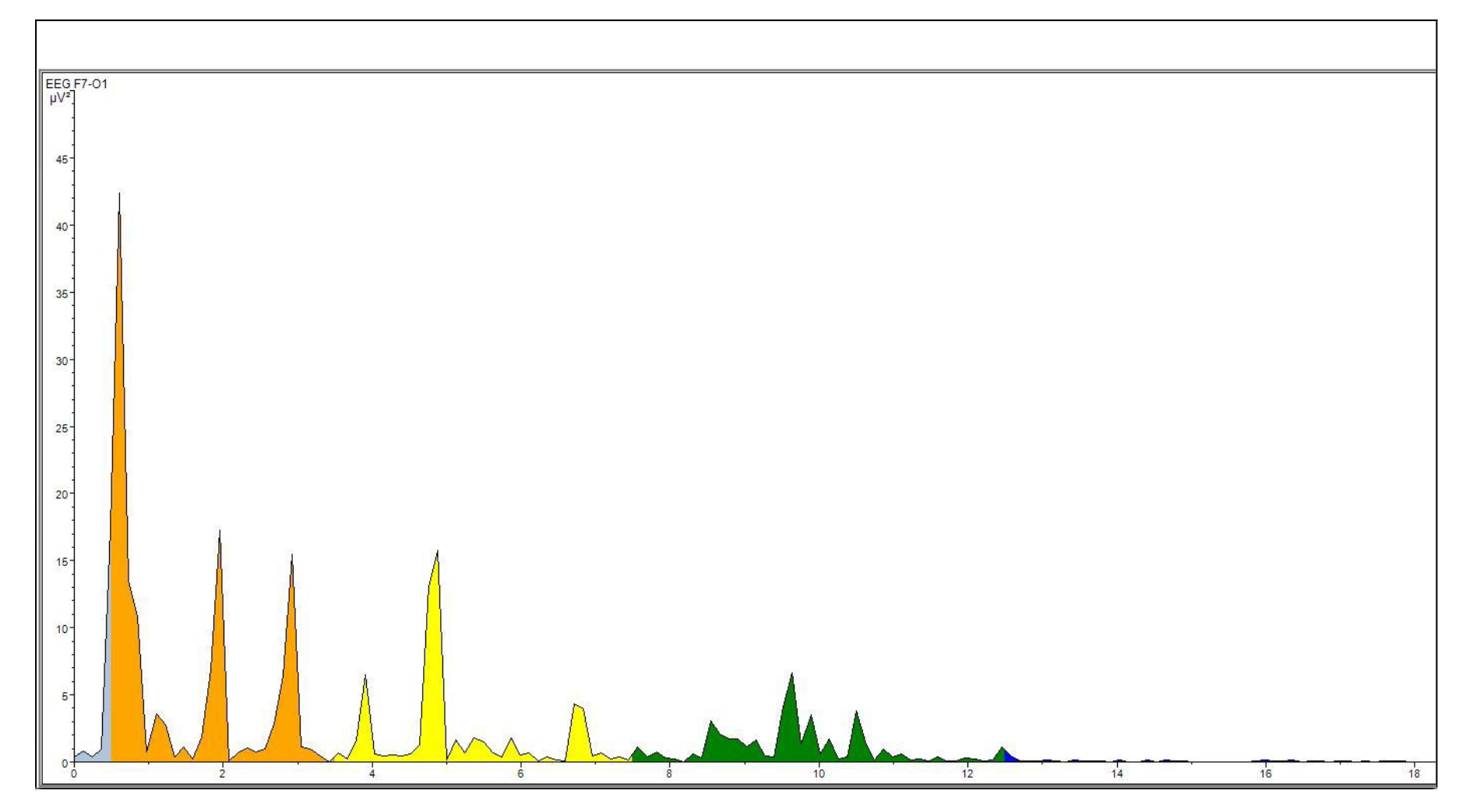
An example FFT produced by BrainVision Analyzer 2 for a 30s epoch of data, in μV^2/Hz. This particular FFT was derived from the F7-O1 channel.

Despite the poor-quality data, we established that delta power can be extracted from the Dreem headband, but additional steps would need to be taken in either a hardware or software domain to ensure that data quality was acceptable.

## DISCUSSION

A key focus for this research was to help inform the potential use of the Dreem 2 headband as a tool to assess sleep in the free-living, ecologically relevant, environment without supervision by the research team. Participants were given initial instruction, both verbal and written, on how to wear the Dreem 2 headband most effectively. We monitored nightly data quality and when a night had less than 70% quality we followed up with a text message to the participant explaining the recording had low quality and we provided basic tips to try and help the participant improve overall data quality. Despite this, quality of recordings was highly variable. 58.5% of all nights were above 70% quality (marked as yellow or green), and 30.7% were above 85% quality (marked as green). Only 145 nights of data were recorded, 51 less than expected for the 14 participants.

Thus, there was 29% missing data prior to any data processing. Nights were lost primarily due to user error by participants, either through an inability to log into the necessary companion Dreem-based app to initiate recording or failure to begin recording during a night. Our findings show it is possible to use the Dreem2 or similar headband device for tracking sleep in the home environment and identify improved data quality as a key need to enhance scientific rigor using such devices. Despite limitations with data quality and missing data, our findings highlight the potential for such devices to help advance the sleep field by providing more detailed sleep tracking data over longer timeframes of weeks to months or even years. Such technology has the potential to help translate data from tightly controlled laboratory trials into real-world settings.

### Bland-Atman Plots

A possible factor in the systemic skew and the widening LOA observed at longer Actiwatch derived sleep durations could be an interplay between the metrics used for determining sleep in the Actiwatch and Dreem 2 headband. The Actiwatch uses primarily motion to assign sleep values, supplemented by user inputs that indicate when the participant is attempting to sleep and wake-up. Alternatively, the Dreem 2 headband quantifies sleep using EEG activity paired with heart rate and motion data. It is possible that in nights with higher sleep opportunities (i.e., time in bed), periods of quiet wakefulness before or after sleep were logged as sleep by the Actiwatch and as wakefulness by the Dreem headband. This hypothesis would align with consistent underreporting of the Dreem relative to the Actiwatch at higher TST. Notably, there are few data points beyond a total sleep time of 480 minutes and several of these data points show significant deviation from the trend. Pairing the Dreem headband with gold-standard PSG would be useful in exploring to what degree the Dreem or Actiwatch truly become less accurate with longer sleep opportunities or extended durations with quiet wakefulness. Because we designed this protocol for the participants to setup and use the Dreem 2 headband on their own in their home environment, the timestamp on the Dreem 2 headbands was not synced with the timestamp on the Actiwatch. This unfortunately prohibits conducting epoch-by-epoch analyses between the actiwatch and Dreem 2 headband in this dataset. Future studies where the timestamps for both devices are synchronized are needed for epoch-by-epoch analyses which will help inform more precisely the scenarios where actigraphy and the Dreem 2 headband are in alignment and disagreement for sleep versus wakefulness.

### Sleep Metrics

First, we observed that stage N3 sleep and REM sleep increased roughly the same amount, on average, during sleep extension versus baseline (19.2 vs 17.9 min, or 26.7% vs 21.7% respectively). The increase in both N3 and REM sleep supports that our participants indeed had insufficient sleep at baseline and show they are physiologically capable of increasing their sleep duration over this four-week sleep extension protocol. Intriguingly, the greatest deficit in most studies with experimental sleep restriction is a reduction in REM sleep, while N3 sleep remains relatively unchanged due to its typical occurrence earlier in the sleep episode (15-17). Therefore, the result that REM sleep increased less than N3 sleep is puzzling, and merits further study in a sleep extension model. If sleep extension and sleep restriction models do not agree, particularly for people with habitual short sleep duration, this may point to a difference of sleep staging or sleep power for people who chose routine short sleep. In other words, there are likely different sleep needs between individuals and understanding individual sleep need could help support better public health recommendations for achieving optimal sleep. N1 and N2 sleep increased as expected based on current sleep restriction models, with N1 representing a brief transitionary period and N2 increasing significantly with extended TST.

Another result of note is the lack of significant differences for SOL, SE, or WASO during sleep extension versus baseline. In the sleep extension segment, a significant drop in SE or a significant increase in SOL or WASO would indicate that extended sleep resulted in a lowered sleep pressure to the degree that sleep quality began to deteriorate. As we did not observe such changes, we can continue to operate under the assumption that the extended sleep is producing longer sleep of similar quality in our participant population. More research is needed to determine if this indicates that extended sleep has health benefits for people with habitual short sleep duration. For example, it is unclear at this time if extending sleep in otherwise healthy young adults who get habitual short sleep duration can help reduce their lifelong risk for chronic disease like diabetes, cardiovascular disease, or Alzheimer’s Disease.

### Delta Power

Our exploration of possible delta power analysis using the data collected by the Dreem 2 headband revealed several significant issues. The automatic Dreem algorithm calculates total recording quality for a single night using data from all channels, such that only a single channel needed to be of high quality for the entire recording to register as high quality. This method of calculating overall night quality creates challenges when trying to restrict channel use to a single spatial area. Because delta power is well documented to vary by cranial region, we focused on two specific channels for our exploratory analyses. Thus, if the only good quality channel was outside our spatial restriction, a night of initially over 85% quality could yield channels of significantly below 70% quality. We observed a possible connection between channel quality and percent data excluded by the BrainVision Analyzer 2 software, suggesting that Dreem assigned quality could have some bearing on actual data usability.

## CONCLUSION

We explored the feasibility of the Dreem 2 Headband for use in at home sleep extension studies. Initially we compared the Dreem 2 headband to actigraphy using a Bland-Altman analysis, where we found significant systemic skew in data with a Dreem assigned quality of lower than 75%. We also established limits of agreement that are narrowest with shortest sleeping times, beginning at roughly 70 minutes above or below actigraphy reported times and widening with increasing sleep duration. We then analyzed sleep metrics for the baseline versus sleep extension conditions, where we observed a significant increase in total sleep time and all sleep phases including N3and REM sleep. We also explored a case study of three individuals with high quality data to determine the feasibility of performing delta power analysis using the Dreem 2 headband. These exploratory analyses found no direct software or hardware obstacles per se. However, data quality declined with the increasing specificity, becoming prohibitively poor when restricting to a single channel. In all, our analyses show the Dreem 2 headband is promising as an at-home sleep monitoring device for sleep research. The primary limiting factor is data quality and this appears at least somewhat dependent on the individual user and highly variable across individuals. With continued work to improve data quality the Dreem has high potential to help advance the sleep field on the whole.

## Data Availability

All data produced in the present study are available upon reasonable request to the authors

